## Supplementary Information for "Strategies to investigate and mitigate collider bias in genetic and Mendelian randomization studies of disease progression"

The two-stage model for LDL-C was univariable only, whereas the two-stage model for HDL-C was additionally run with multivariable adjustment for triglycerides and apolipoprotein B, given expectations of pleiotropy (39, 55). For example, first-stage multivariable models for HDL-C were adjusted for measured triglycerides, the GRS for triglycerides, measured apolipoprotein B, the GRS for apolipoprotein B, age, sex, and GPCs. The second-stage multivariable models for HDL-C were then adjusted for genetically-predicted triglycerides, genetically-predicted apolipoprotein B, age, sex, and GPCs (for the purpose of these models, genetically-predicted triglycerides and genetically-predicted apolipoprotein B were themselves based on multivariable adjustments for HDL-C and apolipoprotein B or triglycerides in first-stage models).

Results of these models supported our expectations of causality, with LDL-C raising the odds of CHD onset, at 1.65 (95% CI = 1.43, 1.91) times higher (**Supplementary Table 1**). In univariable models, higher HDL-C appeared to reduce the odds of CHD onset, at 0.86 (95% CI = 0.77, 0.96) times lower. Upon multivariable adjustment for triglycerides and apolipoprotein B, however, this estimate fully attenuated towards the null, e.g., to an OR of 1.03 (95% CI = 0.94, 1.14) (**Supplementary Table 1**), suggesting that HDL-C does not directly affect CHD onset.

**Supplementary Table 1** One-sample MR estimates of the effect of lipid traits on CHD onset in UK Biobank (N=337,288 eligible)

| | OR (95% CI) for CHD onset per SD higher lipid trait |
|---|---|
| **Positive effect expected** | |
| LDL-C | 1.65 (1.43, 1.91) |
| **Inverse effect expected** | |
| HDL-C, unadj. | 0.86 (0.77, 0.96) |
| **Null effect expected** | |
| HDL-C, adj. for trig. & ApoB | 1.03 (0.94, 1.14) |

Estimates are among adults of European ancestry and are adjusted for age, sex, and the first ten genetic principal components. Models are two-stage prediction substitution regression models with bootstrapped standard errors (100 replications).

**Supplementary Table 2** One-sample MR estimates of the effect of lipid traits on statin use in UK Biobank

| | OR (95% CI) for statin use per SD higher lipid trait | |
|---|---|---|
| | **Among adults overall** N=357,840 eligible | **Among adults with CHD history** N=20,552 eligible |
| LDL-C | 3.85 (3.59, 4.13) | 5.71 (3.62, 9.00) |
| HDL-C | 0.76 (0.73, 0.80) | 0.82 (0.71, 0.95) |

Estimates are among adults of European ancestry and are adjusted for age, sex, and the first ten genetic principal components. Models are two-stage prediction substitution regression models with bootstrapped standard errors (100 replications).
